# The incidence-based dynamic reproduction index: accurate determination, diagnostic sensitivity, and predictive power

**DOI:** 10.1101/2022.04.11.22273599

**Authors:** Robert N. J. Conradt, Stephan Herminghaus

**Affiliations:** Max Planck Institute for Dynamics and Self-Organization (MPI-DS), Am Faßberg 17, D-37077 Göttingen, Germany

## Abstract

Two methods of calculating the reproduction index from daily new infection data are considered, one by using the generation time *t*_*G*_ as a shift (`R`_*G*_), and an incidence-based method directly derived from the differential equation system of an SIR epidemic dynamics model (`R`_*I*_). While the former is shown to have few in common with the true reproduction index, we find that the latter provides a sensitive detection device for intervention effects and other events affecting the epidemic, making it well-suited for diagnostic purposes in policy making. Furthermore, we introduce a similar quantity, 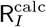, which can be calculated directly from `R`_*G*_. It shows largely the same behaviour as `R`_*I*_, with less fine structure. However, it is accurate in particular in the vicinity of `R` = 1, where accuracy is important for the corrrect prediction of epidemic dynamics. We introduce an entirely new, self-consistent method to derive, from both quantities, an improved 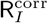 which is both accurate and contains the details of the epidemic spreading dynamics. Hence we obtain `R` accurately from data on daily new infections (incidence) alone. Moreover, by using `R`_*I*_ instead of `R`_*G*_ in plots of `R` versus incidence, orbital trajectories of epidemic waves become visible in a particularly insightful way, demonstrating that the widespread use of only incidence as a diagniostic tool is clearly inappropriate.

PACS numbers:

## I. INTRODUCTION

The outbreak of the illness COVID-19, caused by the SARS-CoV-2 virus, has resulted in a pandemic with unprecedented impact on societies all over Earth. Mitigation measures included complete lockdowns of societal life, with severe social, economic, and individual consequences [1, 2]. The dramatically varying success [3–6] of the interventions owed in part to cultural differences [3], but also to only limited understanding of infection spreading dynamics and a severe lack of established methods in epidemic state diagnosis and prediction. Improvement of this general situation is imperative, in particular as similar events are expected to strike more often in the future [7–9].

In search for optimized strategies, a two-fold view must be adopted. One the one hand, one needs to understand, in retrospect, which interventions have had what effect on the epidemic spreading dynamics, in order to properly design future interventions. This requires sensitive diagnosis tools for assessing the state of the epidemic on a (if possible) daily basis. On the other hand, tools are needed for predicting the future of epidemic dynamics as reliably as possible if conditions are known. Aside from extensive simulation, this requires careful analysis of data, such as the number of infected citizens [2, 10].

Here we discuss the system in terms of an SIR model [11, 12], referring to the number of susceptible (S), infected (I), and recovered (R) individuals, respectively, in a population of *N* citizens. Here we identify with *R* all those who are neither susceptible nor infected (*R* = *N − S − I*), which includes those who are deceased. While the ratio of deceased vs. recovered individuals is of greatest societal concern, it can be disregarded here, as we will solely discuss prevention measures addressing the spreading of the disease. We define *I* as the number of individuals who carry sufficient viral load to be contagious. They are assumed to remain in this state for an average duration *τ*. Although the viral load changes with time during the illness, contagion can be sufficiently well described by this simple picture for our purposes [13].

The spreading dynamics of an epidemic can then be described by a set of two equations [12],

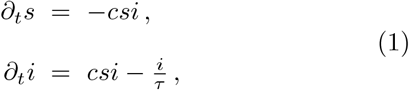

where *∂*_*t*_ is the derivative with respect to time, while *s*(*t*) = *S/N* and *i*(*t*) = *I/N* are the fractions of susceptible and infected individuals in the population, respectively. The constant *c* is the average number of new infections a single infected individual would cause per unit time in an otherwise infection-free (but susceptible) population. It is accessible to interventions such as closing schools, wearing facial masks etc., but this shall not concern us here, as we focus solely on methods to detect the current stage of an epidemic and to predict its near future development, at given *c*.

## II. THE REPRODUCTION INDEX

The base reproduction index, `R`_0_, is related to *c* via

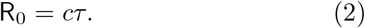

It denotes the total average number of individuals newly infected by a single infected one under the above conditions. Since the probability of infection is directly proportional to the fraction of susceptibles, we have

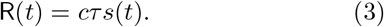

for the dynamic reproduction index, `R`(*t*) [12]. The latter is of tremendous importance for assessing the current status of an epidemic. If `R` = 1, *I* stays constant, but when `R` *>* 1, each infected individual causes more than one new infection on average, such that *I*(*t*) increases exponentially. It is thus of major interest to determine `R` from epidemiological data as accurately as possible, in particular in the vicinity of unity.

By combining eq. (3) with the first eq. (1), we obtain

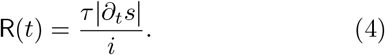

This relies only on quantities which can be derived from data usually available from the health care system. *∂*_*t*_*S* represents the number of new infections per unit time (daily incidence) and can be considered known accurately. *τ* is known from clinical experience with the disease, and *I*(*t*) (and hence *i*) can be estimated once *τ* is known. We will see below that it is in particular the dynamic variations of `R` which yield considerable insight into the infection process.

Since infection data are discrete data collected on a daily basis [14], we will now write down a discrete version of eq. (4). The daily incidence will be called 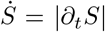. *τ* as well as *t* will henceforth be expressed in units of days, and treated as discrete variables. We then may be tempted to simply write 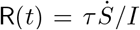. However, we must be aware that for rather general infrastructural reasons, the reporting efficiency of infection numbers varies characteristically, e.g., on weekends. We therefore should provide for suitable averaging. Hence we write

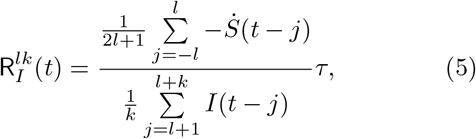

where *l* and *k* are parameters determining the intervals over which 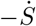 and *I* are being averaged, respectively. Since the typical variability of data reflects the sequence of seven weekdays, it appears reasonable to average 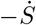 over seven consecutive days. If we furthermore average the (less variable) number of infected individuals over a period *k* = *τ*, we obtain

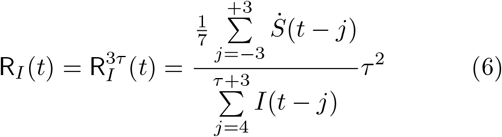

for the incidence-based dynamic reproduction index. While 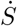 is known precisely, *I* can only be estimated based, among others, on *τ*. However, this has only minor effects on the accuracy of `R`_*I*_ and its dynamic variations, as *I* enters only as an average over the duration *τ*.

The merits of `R`_*I*_ show up clearly when compared to other definitions of reproduction indices which are cur-rently used in epidemic data based diagnostics of the infection dynamics. Since we will later use data from Germany in our analysis, we refer to what is issued by the Robert Koch Institute (RKI) in Germany as the “reproduction index”. It is based on the idea of calculating the ratio of incidence data, taken on two successive instants, separated by a delay time *t*_*G*_ [15]. The latter is called the generation time and represents the average time interval between an infection and a subsequent “successful” transmission of the infection to a third person. The RKI uses *t*_*G*_ = 4 days. The definition of this generation-time based reproduction index is then [16, 17]

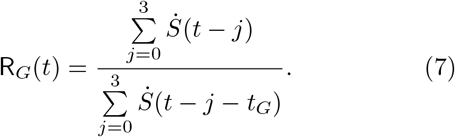

Its mathematical meaning becomes clearer in a continuous formulation,

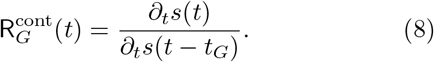

This can be written as

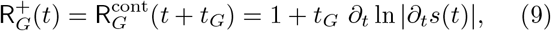

where we have truncated the Taylor expansion after the first term. This reveals that `R`_*G*_ is directly related to the logarithmic derivative of the daily incidence, |*∂*_*t*_*s*(*t*) |, with some delay equal to *t*_*G*_. Hence whenever the daily incidence happens to vary exponentially, *∂*_*t*_*s*(*t*) ∝exp (`R`−1)*t*, `R`_*G*_ can indeed be interpreted as a reproduction index. At any other time, however, when this is not the case, the use of `R`_*G*_ as a reproduction index lacks mathematical foundation. Note furthermore that while *c* enters directly in `R` (as given by eqs. (3) and (4)), it cancels out in all expressions for `R`_*G*_. Hence public measures affecting *c* will readily show up in `R`_*I*_, which has been defined according to eq. (4), but not in `R`_*G*_.

## III. APPLICATION TO EPIDEMIC DATA

From data obtained in Germany during the SARS-CoV2 pandemic in 2020 and 2021 [18], let us now calculate `R`_*I*_ (*t*) and `R`_*G*_(*t*) by means of eqs. (6) and (7), respectively. The result is displayed in the top panel of Fig. 1, exposing the remarkable differences between the two quantities. There is a strong tendency of `R`_*G*_(*t*) to stay closer to unity than `R`_*I*_ (*t*), which reflects the dynamics more pronouncedly. Events like the strong increase of `R`_*I*_ (*t*) up to a value of 2.3 at the end of October 2020 (around day 245) hardly show up in `R`_*G*_(*t*).

**FIG. 1:**
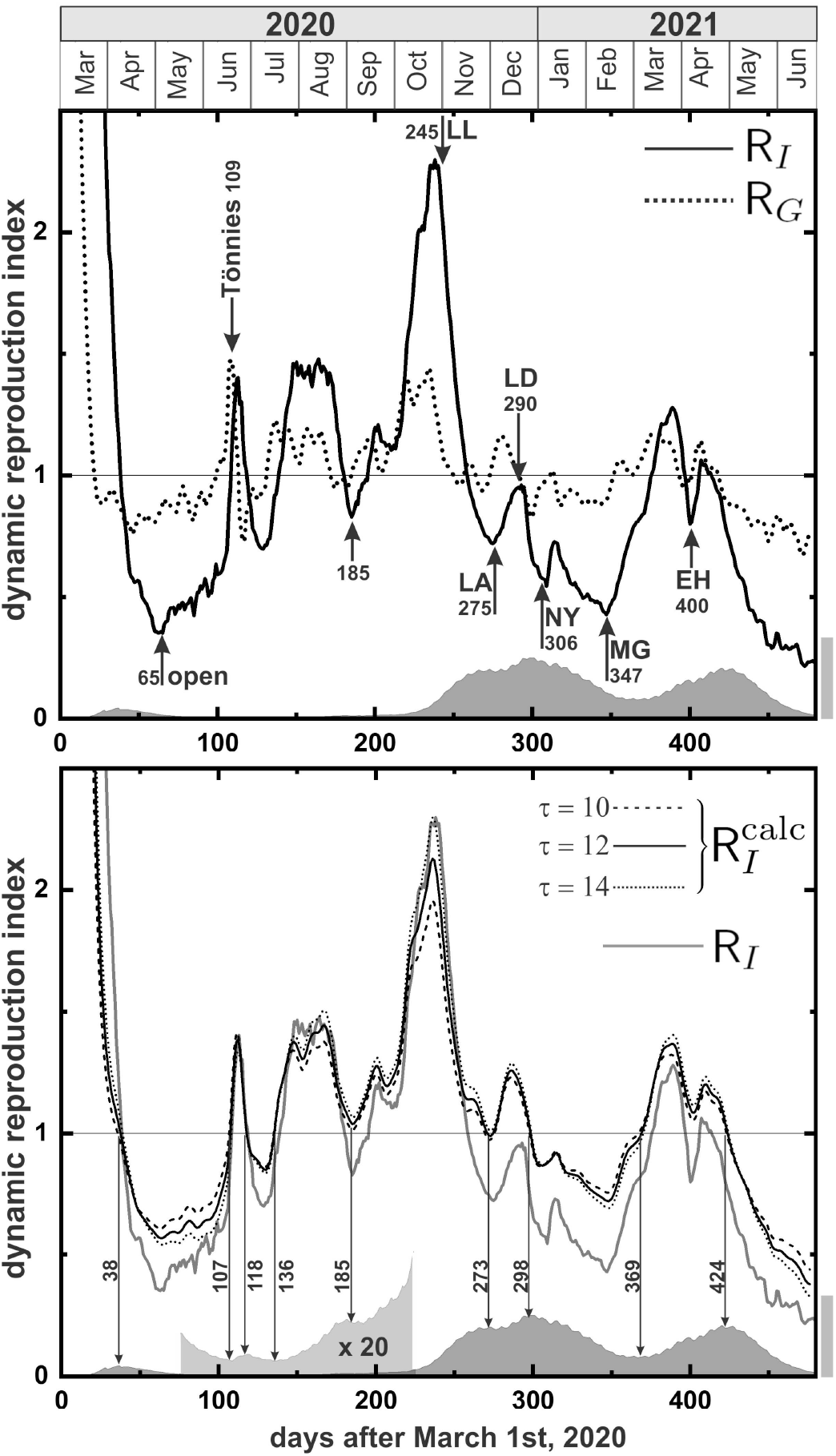
*Top panel:* the two reproduction indices `R`_*G*_(*t*) (dotted) and `R`_*I*_ (*t*) (solid) as obtained form infection data in Germany during the SARS-CoV2 pandemic. The arrows indicate certain events and interventions (see table I) as discussed in the text. The grey shaded curve at the bottom indicates the prevalence (number of infected). the corresponding grey scale bar to the right corresponds to one half million people. *Bottom panel:* `R`_*I*_ (*t*) as obtained from `R`_*G*_(*t*) through eq. (14) for different values of *τ* (black curves) and numerically from infection data (grey, same as black curve in top panel). The vertical arrows are located where the black curves reach unity, and coincide well with the extrema of the prevalence. In the interval *t* ∈ [75, 225], the prevalence curve has been scaled by a factor of 20 for visibility.

For a more detailed discussion, a number of important events are listed in table I. After day 63, which corresponds to May 5, 2020, we see a sharp increase of `R`_*I*_. At the end of the first wave during March and April, the German Chancellor and the Conference of prime ministers of the Länder had decided to relax public life to almost normal conditions. Hence stores, restaurants, cultural institutions, and museums were opened. Because there was only little change in incidence (and prevalence) during the rest of spring and summer, it went unnoticed that the reproduction index was undergoing strong changes. That these were much less pronounced in `R`_*G*_ (which was used) than in `R`_*I*_ made their detection particularly difficult.

**TABLE I:** A number of events marked in the top panel of Fig. 1.

| t | date | label | event |
| --- | --- | --- | --- |
| [d] | (d/m/y) | (Fig. 1) |  |
| 65 | 5/5/2020 | open | reopening stores and restaurants |
| 109 | 18/6/2020 | Tönnies | outbreak at Tönnies company site (Gütersloh) is noticed |
| 245 | 1/11/2020 | LL | lockdown light |
| 275 | 1/12/2020 | LA | december lockdown announced |
| 290 | 16/12/2020 | LD | december lockdown starts |
| 306 | 1/1/2021 | NY | new year's day |
| 347 | 11/2/2021 | MG | Mardi Gras |
| 400 | 5/4/2021 | EH | Easter holidays |

A few days after a strong rise in `R`_*G*_ was noticed, a major disease outbreak was reported on day 109 at the Tönnies sloughterhouse site near Guütersloh, among the large number of loan workers living at the site. But since prevalence remained low after that, the further rise of `R` was not noticed, or taken seriously, certainly in part due to the noisiness of the data. After the incidence then rose very sharply in the fall of 2020, the German government decided on what became known as a “lockdown light”, starting from November 2nd (day 245). This was associated with a hope of possibly easing restrictions for Christmas.

Because the pronounced decrease of the reproduction index is not reflected by `R`_*G*_, and incidence did not seem to decrease, it was not noticed that the situation was actually relaxing during November. Consequently, a hard lockdown (including widespread closures of shops, businesses, schools, etc.) was announced on December 1st (day 275), which would start on December 16th (day 290). In particular, the relaxations for Christmas that had been previously promised were withdrawn. This announcement led to a short-lived but sharp increase in infections, as many people squeezed through the two-weeks bottleneck for their Christmas shopping. The subsequent decline (presumably due to the start of the Christmas school holidays) abruptly terminated on new year’s day (day 347), when many people had visited relatives and friends. A similar feature appears on day 400 at the Easter holidays, for similar reasons. Clearly, most (if not all) of these features are only poorly (if at all) discernible in `R`_*G*_.

Nevertheless, it may seem that some features in `R`_*G*_(*t*) appear a little earlier than corresponding features in `R`_*I*_ (*t*). This is apparent most clearly from the points where unity is crossed, which for `R`_*G*_(*t*) lie significantly before those for `R`_*I*_ (*t*). This could be interpreted as `R`_*G*_ being better suited for forecast purposes than `R`_*I*_, at it discloses the same information at an earlier time. However, this turns out to be a delusive mathematical artifact. As we see from eq. (9), with help form eq. (1), 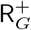 can be written as

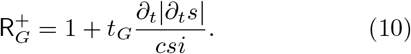

By means of eqs. (4) and (1), the numerator of the second term can be expressed as

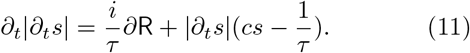

Consequently, we have

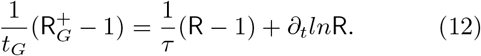

In other words, the deviation of 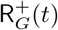 from unity is composed of a term proportional to the deviation of `R`(*t*) from unity and the logarithmic time derivative of `R`(*t*). Hence `R`_*G*_ has a much more complex structure than `R`, exhibiting additional features (and additional noisiness) from the time derivative of `R`. It hence cannot be interpreted in terms of a true reproduction index.

We have seen above that `R`_*I*_ is a powerful diagnosis tool, as it reacts sensitively to events and interventions in society. Nevertheless, it comes with its downsides. From the second wave (around day 300) we see that the absolute magnitude of `R`_*I*_ cannot be accurate, as the number of infections rises considerably shortly before day 300, while `R`_*I*_ is clearly smaller than unity. This may be attributed to the fact that `R`_*I*_ depends on *τ* (cf. eqs. (6)), which cancels out for `R`_*G*_, as it is obvious form eq. (7). The inherent problem is that *τ* may not only be known with poor accuracy, but it can also undergo gradual changes during the epidemic. An obvious cause may be the gradual appearance of mutations, which often result in shifts in the clinical picture, possibly including changes in the duration of the illness, hence in *τ*. It will only rarely be possible to receive regular reliable data on *τ*.

## IV. SELF-CONSISTENT CORRECTION OF R

It turns out that the independence of `R`_*G*_ from *τ* may be exploited here by inverting eq. (12), in order to calculate `R`_*I*_ from `R`_*G*_. If we denote by 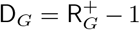 the deviation of 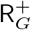 from unity, we can rewrite eq. (12) (by multiplying with *τ* and dividing by `R`) into

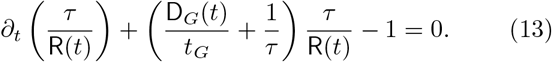

This is a linear differential equation in *τ/*`R` and can be solved by means of the method of variation of parameters.

The result is

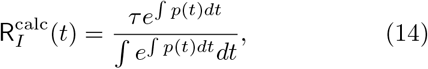

where

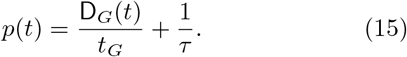

One finds that 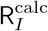 depends on *τ* only weakly, because its role as a prefactor im eq. (14) and its appearance in *p*(*t*) cancel each other to a large extent. In the bottom panel of Fig. 1, 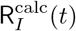 thus obtained is plotted for three different values of *τ* as the black curves. The grey curve is `R`_*I*_ as in the top panel. Clearly, all of the more prominent features of `R`_*I*_ are reproduced.

There are two main differences between 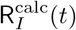 and `R`_*I*_ (*t*). First, there is a vertical shift which varies with time only very slowly. Second, much finer details are visible in `R`_*I*_. The striking feature of 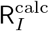 is that close to its transitions through unity, there is almost no sensitivity to *τ*. Hence should *τ* vary over the course of the epidemic by, e.g., as much as 40 percent (as between the dashed and the dotted curve), the shape of 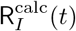 would not change much. In particular, the points where it hits unity do not change their position appreciably. As the vertical arrows show, these points are very close to the extrema of the number of infected people, as one would correctly expect for the reproduction index.

Hence what we display in the bottom panel of Fig. 1 may well be called the best of both worlds. In `R`_*I*_ (*t*) we see very fine details which allow to identify the effects of social events, and to assess the effectiveness of public interventions in retrospect. In 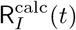, which we derived from `R`_*G*_(*t*) by means of eq. (14), we see less detail, but obtain a more accurate estimate of the reproduction index. This allows for more reliable predictions of near-future epidemic dynamics, as 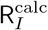 is particularly accurate close to unity. The difference between `R`_*I*_ and `R`_*G*_ is presumably due to a (slowly) varying *τ*.

If we explicitly demand variation in *τ* to be slow as compared to the rapid variations seen in `R`_*I*_, we can approximately determine *τ* (*t*) from `R`_*I*_ and `R`_*G*_ in a self-consistent manner. First we calculate the ratio 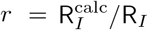 and fit a low order polynomial *q*(*t*) to it, such that high-frequency components are cut off. Then we set *τ*_1_(*t*) = *τ*_0_*q*(*t*), where *τ*_0_ is the constant *τ* we used initially calculating `R`_*I*_ from eq. (6). This yields a new 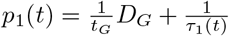, which we use, together with *τ*_1_(*t*), to recalculate 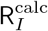 from eq. (14). This is repeated until the result of the polynomial fit has become stable. 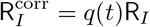 is then a reproduction index which is self-consistently matched to the one obtained from `R`_*G*_ and should, just as the latter, provide high accuracy (in particular close to unity) while keeping all the fine details we found to be present in `R`_*I*_, which we initially calculated from eq. (6).

The result we obtained using fourth-order polynomials for *q*(*t*) is presented in Fig. 2. After five iterations, the result for *q*(*t*), did not change anymore. As one can clearly see, 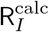 has been rather well matched to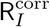, aside from some high-frequency variations. We have exploited the separation of time scales between *τ* and `R`_*I*_. One may admit faster variations to *τ* in order to achieve an even better match between 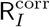 and 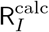, but this shall not concern us here. *q*(*t*) is indicated by the dotted curve in the figure, to be read off the left scale. The scale to the right shows the corresponding values of *τ*. Note that the pronounced peak around day 240 reaches a very high value, deviating from unity by about four times more strongly than `R`_*G*_ (dotted curve in Fig. 1). This highlights the importance of our method to calculate `R`_*I*_ for predictions of near-future epidemic spreading dynamics. Around day 240, using `R`_*G*_ would have (or actually has) over-estimated the doubling time of infections by a factor of 4.5.

**FIG. 2:**
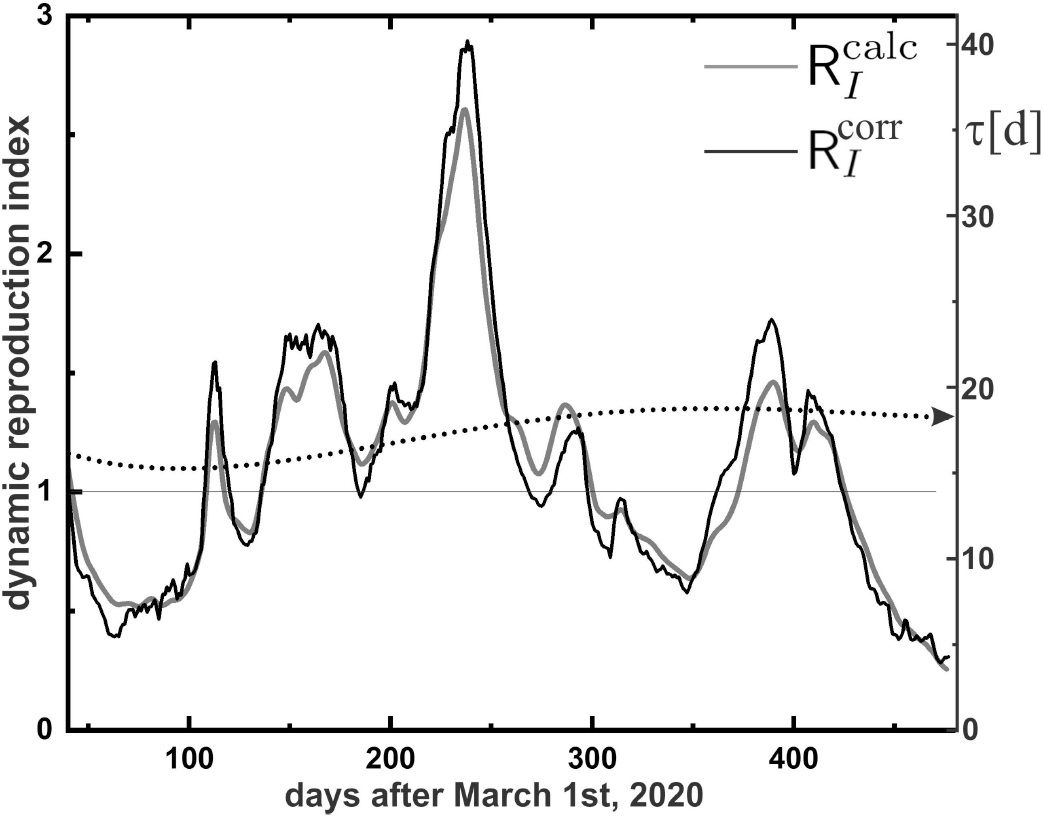
Self-consistent matching of the low-frequency components of `R`_*I*_ (*t*) (black) and 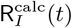 (grey). Matching has been achieved by setting *τ* = *q*(*t*)*τ*_0_, where *τ*_0_ = 14 has been used for the initial calculation of `R`_*I*_ (*t*), and *q*(*t*) is a fourth-order polynomial. It is indicated by the dotted curve. The corresponding values of *τ* (*t*) can be read off the scale to the right.

## V. REVEALING THE ORBITAL STRUCTURE OF EPIDEMIC WAVES

Finally, it is instructive to elaborate on some additional aspects of data presentation and analysis. In Fig. 3a we plot the number of new infections during a period *τ* against the prevalence, i.e., the total number of currently ill individuals. The ordinate is calculated from the seven-day averaged incidence by multiplying with *τ/*7. Each of the small circles represents one day, with the symbol style representing the three epidemic waves (open, first wave. full grey, second wave. full black, third wave). The data are gathering into an elongated cloud along the first diagonal (dashed line). If we assume that about 3% of individuals infected with SARS-CoV2 need intensive care, we can estimate the maximum prevalence the society could bear. Since there are 16734 intensive care beds in Germany [19], we conclude that the displayed range of the abscissa represents the maximum “acceptable” range of prevalence (one half million infected individuals).

**FIG. 3:**
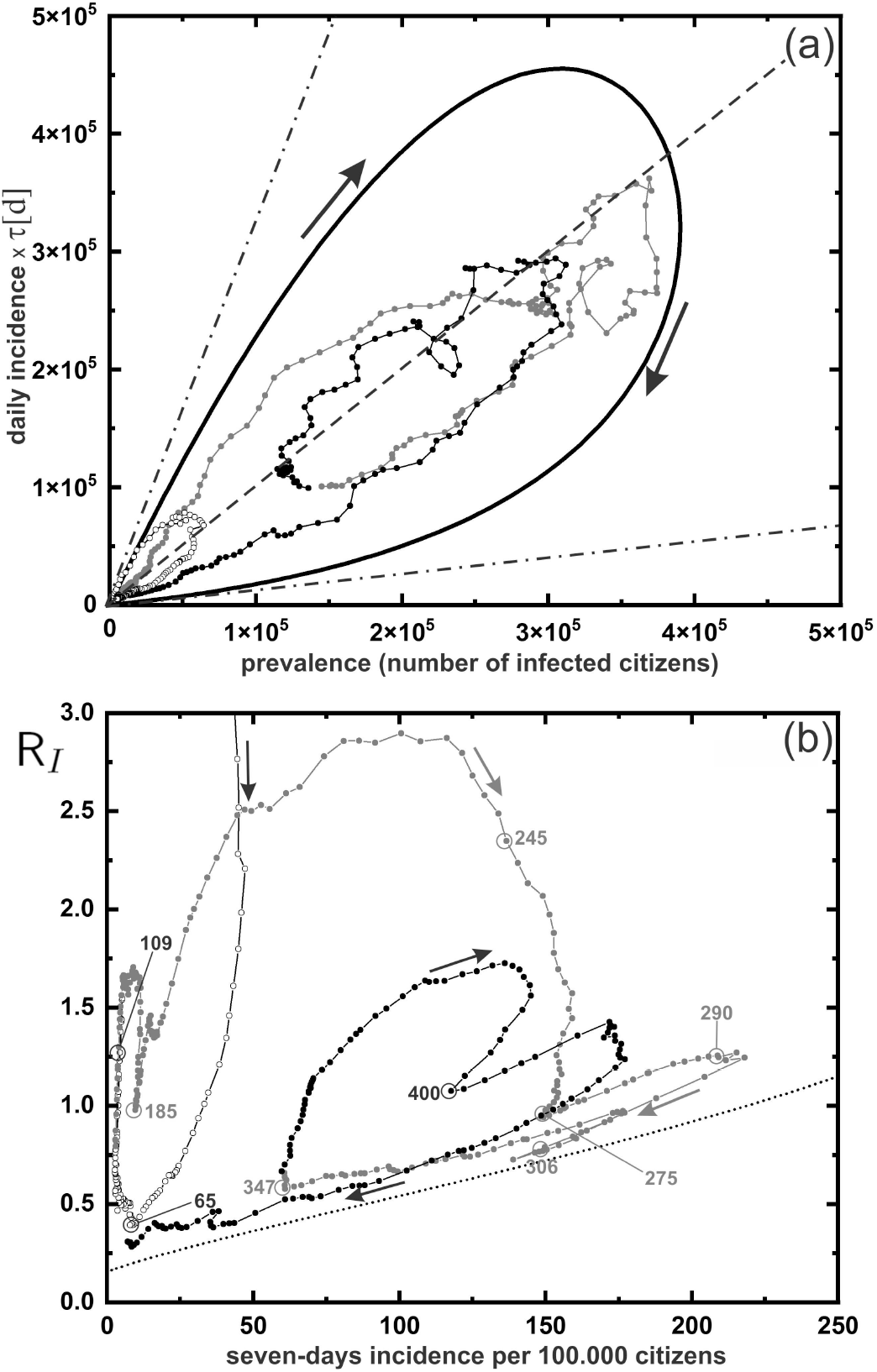
Day-by-day epidemic trajectories (circles with polygons). First wave (open), *t* < 130. Second wave (filled grey), *t* ∈ [130, 351]. Third wave (filled black), *t* > 351. (a) Total number of new infections within a time interval *τ* versus total number of acutely infected. Each wave appears as a clockwise orbit, with smaller sub-orbits. Data points tend to group along the first diagonal (dashed line). Solid curve: a sample simulation of an epidemic trajectory, showig the generic clockwise orbit structure, very similar to the data from the first wave. Both are asymptotic to the dash-dotted line with slope `R`_0_ = 3.3. The second (lower) asymptote corresponds to the terminal value of *R* = 0.14. The horizontal axis spans the maximum range of SARS-CoV2 infections acceptable to the German health system (about one half million). (b) Reproduction index versus seven-days incidence. The orbit structure is more clearly revealed. This presentation allows to assess in which phase of an epidemic wave the system currently is. All data stay just above the asymptotic (dotted curve) solution of eq. (1), which enters the vertical axis at the terminal value of *R*_1_ = 0.14. Various incidents mentions in table I are marked along the trajectory. Note that `R` is generally a forerunner to incidence, and a new orbit is marked as a sharp increase of `R`. Hence one may miss important developments when monitoring incidence alone.

In order to analyse the internal structure of the data cloud, we compare with a numerically simulated sample trajectory of eq. (1), which is shown as the solid curve. It forms a lobe, starting off at the origin with a slope equal to `R`_0_, proceeds clockwise (arrows) and re-enters the origin at an inferior slope of `R`_1_ = 0.14 (lower dash-dotted line) for *t→∞*. The initial slope, which is indicated by the upper dash-dotted line, follows eq. (4), since the ordinate and abscissa just represent *τ* |*∂*_*t*_*s*| and *i*, respectively. For the simulation, we have set `R`_0_ = 3.3 in order to match typical values assumed for SARS-CoV2. The size of the lobe corresponds to the number of people affected by the epidemic. As a consequence of the structure of the solutions to eq. (1), `R`_1_ is a function of `R`_0_, and `R`_1_(3.3) = 0.14.

In fact, the data representing the first wave (open circles close to the origin) exhibit just the same lobe shape, initially following the dash-dotted line, and as time proceeds is traversed in clockwise direction. A closer look at the second (grey) and third (black) wave reveals that their trajectories tend to form clockwise orbits as well, with smaller sub-orbits, thus exposing additional fine structure of the infection dynamics. Positioning data within the course of an orbit may be very useful to assess the stage of the epidemic spreading.

This orbit structure is revealed even more clearly in a different presentation, when we plot `R`_*I*_ versus incidence, as shown in Fig. 3b. What we can clearly see, for instance, is that the situation was already on a relaxing path when the “lockdown light” came into effect on day 245. When the hard December lockdown was announced on day 275, the orbit was in fact almost finished and back towards the origin. Things would thus have probably eased off by themselves, if no major mistakes would have been made. After Mardi Gras (day 347), incidence stayed calm, and `R`_*G*_ remained featureless within noise level, as Fig. 1 (top) shows. Hence no countermeasures were taken. The trajectory in Fig. 3b, however, clearly shows that this was when a new orbit had formed. This very probably launched the third wave. Most importantly, we see from Fig. 3b that the widespread exclusive use of the incidence (abscissa) for assessing the state of the epidemic is void of any sound basis, as it does not reflect the orbital structure of epidemic waves. In particular, the use of threshold values for incidence in legislation on public mitigation interventions is clearly not appropriate.

## VI. CONCLUSIONS

We have presented analysis tools for different aspects of epidemic mitigation and management interventions. First, we showed that from data of daily incidence (and prevalence derived therefrom) one can derive, via eq. (6), an accurate value for the reproduction index. `R`_*I*_ (*t*) provides a very sensitive seismograph of the currrent state of the epidemic. Second, we have shown that a similar quantity, 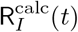, can be derived via eq. (14) from the generation-based reproduction index, `R`_*G*_. It has the same general behaviour as `R`_*I*_ (*t*), but has the particular merit of being very accurate whenever it is close to unity. This is of great interest for forecasting the epidemic development, which abruptly changes at `R` = 1. The fact that it lacks much of the fine details showing up in `R`_*I*_ (*t*) makes it particularly suited for polynomial extrapolation towards unity. Third, we have shown that a much improved (corrected) form of `R`(*t*) can be obtained in a self-consistent manner, thereby also providing the so far only poorly known, slowly varying, *τ* (*t*). Finally, we have demonstrated that a presentation of standard epidemic data in the plane spanned by the reproduction index and the incidence exploits the internal orbit structure of epidemic waves in a way beneficial for assessing the current epidemic state of affairs. It should be a very useful tool for policy makers during dangerous epidemics, such as the recent outbreak of COVID-19.

## Data Availability

All data are available under "daily reports of the Robert-Koch-Institute / Germany"

https://www.rki.de/EN/Home/homepage_node.html;jsessionid=6B1BE151A64136D8C071E6DF5EF019DC.internet082

## VII. ACKNOWLEDGEMENTS

The authors are grateful to Paul Leiderer, Christoph Klein, and Armin Lambacher for inspiring discussions and critical reading of the manuscript. This paper is dedicated to all those who died in the Covid19 pandemic, especially to those who passed away in loneliness.

